# Safety and Immunogenicity of an Inactivated Recombinant Newcastle Disease Virus Vaccine Expressing SARS-CoV-2 Spike: Interim Results of a Randomised, Placebo-Controlled, Phase 1/2 Trial

**DOI:** 10.1101/2021.09.17.21263758

**Authors:** Punnee Pitisuttithum, Viravarn Luvira, Saranath Lawpoolsri, Sant Muangnoicharoen, Supitcha Kamolratanakul, Chaisith Sivakorn, Piengthong Narakorn, Somchaiya Surichan, Sumalee Prangpratanporn, Suttida Puksuriwong, Steven Lamola, Laina D Mercer, Rama Raghunandan, Weina Sun, Yonghong Liu, Juan Manuel Carreño, Rami Scharf, Weerapong Phumratanaprapin, Fatima Amanat, Luc Gagnon, Ching-Lin Hsieh, Ruangchai Kaweepornpoj, Sarwat Khan, Manjari Lal, Stephen McCroskery, Jason McLellan, Ignacio Mena, Marcia Meseck, Benjaluck Phonrat, Yupa Sabmee, Ratsamikorn Singchareon, Stefan Slamanig, Nava Suthepakul, Johnstone Tcheou, Narumon Thantamnu, Sompone Theerasurakarn, Steven Tran, Thanakrit Vilasmongkolchai, Jessica A White, Adolfo Garcia-Sastre, Peter Palese, Florian Krammer, Kittisak Poopipatpol, Ponthip Wirachwong, Richard Hjorth, Bruce L Innis

## Abstract

**Background:** Production of affordable coronavirus disease 2019 (COVID-19) vaccines in low- and middle-income countries is needed. NDV-HXP-S is an inactivated egg-based Newcastle disease virus vaccine expressing the spike protein of severe acute respiratory syndrome coronavirus 2 (SARS-CoV-2). It’s being developed in Thailand, Vietnam, and Brazil; herein are initial results from Thailand.

**Methods:** This phase 1 stage of a randomised, dose-escalation, observer-blind, placebo-controlled, phase 1/2 trial was conducted at the Vaccine Trial Centre, Mahidol University (Bangkok). Healthy adults aged 18-59 years, non-pregnant and negative for SARS-CoV-2 antibodies were eligible. Participants were block randomised to receive one of six treatments by intramuscular injection twice, 28 days apart: 1 µg±CpG1018 (a toll-like receptor 9 agonist), 3 µg±CpG1018, 10 µg, or placebo. Participants and personnel assessing outcomes were masked to treatment. The primary outcomes were solicited and spontaneously reported adverse events (AEs) during 7 and 28 days after each vaccination, respectively. Secondary outcomes were immunogenicity measures (anti-S IgG and pseudotyped virus neutralisation). An interim analysis assessed safety at day 57 in treatment-exposed individuals and immunogenicity through day 43 per protocol. ClinicalTrials.gov (NCT04764422).

**Findings:** Between March 20 and April 23, 2021, 377 individuals were screened and 210 were enrolled (35 per group); all received dose one; five missed dose two. The most common solicited AEs among vaccinees, all predominantly mild, were injection site pain (<63%), fatigue (<35%), headache (<32%), and myalgia (<32%). The proportion reporting a vaccine-related AE ranged from 5·7% to 17·1% among vaccine groups and was 2·9% in controls; there was no vaccine-related serious adverse event. The 10 µg formulation’s immunogenicity ranked best, followed by 3 µg+CpG1018, 3 µg, 1 µg+CpG1018, and 1 µg formulations. On day 43, the geometric mean concentrations of 50% neutralising antibody ranged from 122·23 IU/mL (1 µg, 95% CI 86·40-172·91) to 474·35 IU/mL (10 µg, 95% CI 320·90-701·19), with 93·9% to 100% of vaccine groups attaining a ≥4-fold increase over baseline.

**Interpretation:** NDV-HXP-S had an acceptable safety profile and potent immunogenicity. The 3 µg and 3 µg+CpG1018 formulations advanced to phase 2.

**Funding:** National Vaccine Institute (Thailand), National Research Council (Thailand), Bill & Melinda Gates Foundation, National Institutes of Health (USA)

## Introduction

There remains a shocking imbalance in the global distribution of coronavirus disease 2019 (COVID-19) vaccines.^1^ To achieve control of the COVID-19 pandemic in low- and middle-income countries (LMICs) where most of the global population resides, there must be a great increase in sustainable supply of affordable vaccines. The manufacturing capacity for egg-based inactivated influenza vaccines (IIV) is among the largest in the world; these facilities, some in middle-income countries and operating for less than six months per year, use locally produced embryonated eggs to make more than a billion doses annually of affordable human vaccines.^2^ To enable these manufacturers to respond to the COVID-19 pandemic, we developed a COVID-19 vaccine for production in eggs, based on a Newcastle disease virus (NDV) expressing the ectodomain of a novel membrane-anchored, prefusion-stabilized severe acute respiratory syndrome coronavirus 2 (SARS-CoV-2) spike protein construct, wherein virions are purified and inactivated (NDV-HXP-S).^3-5^

From September to November 2020, manufacturers in Thailand, Vietnam, and Brazil modified their IIV manufacturing process to optimize production of beta-propiolactone (BPL)-inactivated NDV-HXP-S, achieving high yields at pilot scale; the result was three similar processes. A preclinical evaluation of their vaccine candidates, formulated with and without CpG1018, a toll-like receptor 9 agonist adjuvant (Dynavax Technologies)^6^ confirmed that they were highly immunogenic and protective in hamsters^5^ with no sign of toxicity in rats at the maximum human doses planned for evaluation (3 µg S protein+1·5 mg CpG1018; 10 µg S protein). These results enabled all three manufacturers to initiate clinical development of their vaccine candidates. Herein, we report interim safety and immunogenicity data generated in the phase 1 portion of an adaptive phase 1/2 clinical trial evaluating the NDV-HXP-S vaccine candidate developed by The Government Pharmaceutical Organization of Thailand (GPO). These results provide the first evidence in humans that the NDV vector technology expressing a six-proline prefusion-stabilized spike protein construct offers a unique platform for affordable manufacturing of a well-tolerated and highly immunogenic COVID-19 vaccine.

## Methods

### Study design and participants

The phase 1 segment of a randomised, observer-blind, placebo-controlled, phase 1/2 trial was conducted at the Vaccine Trial Centre, Faculty of Tropical Medicine, Mahidol University (Bangkok, Thailand). Participants were recruited from individuals known to the Centre and through advertisements. Healthy adults 18–59 years of age with body mass index 17 to 40 kg/m^2^,-negative hepatitis B surface antigen and SARS-CoV-2, HIV, and hepatitis C antibody tests were eligible to participate. A negative urinary pregnancy test was required of women having reproductive capacity prior to administration of each study vaccine dose. Complete eligibility criteria are described in the trial protocol provided in the supplementary materials.

Written informed consent was obtained from all participants. The trial complied with the Declaration of Helsinki and Good Clinical Practice. This study was approved by the Ethics Committee of the Faculty of Tropical Medicine, Mahidol University (TMEC 21-005) and authorized by the Thailand Food and Drug Administration (FDA-21-018).

### Randomisation and masking

Enrolled subjects were randomly assigned in sequence to one of 6 equal groups (vaccine containing 1 µg S ± 1·5mg CpG1018 adjuvant, 3 µg S ± 1·5mg CpG1018 adjuvant, 10 µg S, or saline placebo). Subjects were enrolled in stages, each including active treatment and placebo groups, using a computer-generated block randomisation sequence prepared by an independent statistician; an unblinded pharmacist team dispensed each treatment according to the randomization sequence. The first 18 subjects (sentinel cohort) were enrolled to three sequential sentinel groups; 3:1, 1 µg and placebo; 3:3:1, 3 µg or 1 µg+CpG1018 and placebo; and 3:3:1, 10 µg or 3 µg+CpG1018 and placebo, After safety data were reviewed for the sentinel groups, the next 192 subjects were randomised in 5-dose-cohorts; 32:6, 1 µg and placebo; 32:6, 3 µg and placebo; 32:6, 1 µg+CpG1018 and placebo; 32:7, 10 µg and placebo; and 32:7, 3 µg+CpG1018 and placebo. All participants and personnel other than the unmasked pharmacy team and vaccinators were masked to treatment.

### Procedures

The recombinant NDV-HXP-S vaccine was manufactured according to current Good Manufacturing Practice by the GPO in their Influenza Vaccine Plant (Saraburi, Thailand) using locally procured embryonated eggs inoculated with a master virus seed made and extensively tested for adventitious agents by the Icahn School of Medicine at Mount Sinai (New York, USA). After incubation for 72 hours at 37°C, eggs were chilled overnight at 4°C, then the allantoic fluids were harvested, clarified, and concentrated. Recombinant virus particles were purified from the concentrated harvest by two sequential continuous flow sucrose gradient centrifugations, diafiltered against phosphate-buffered saline (PBS), inactivated by treatment with 1:4000 BPL for 24 hours at 4°C, and 0.2 micron filter-sterilized. Vaccine potency was measured by direct enzyme-linked immunosorbent assay (ELISA) using a human monoclonal antibody (CR3022^7^) to SARS-CoV-2 spike glycoprotein S1 (LakePharma Inc) and an NDV-HXP-S standard that had been calibrated to a purified HXP-S reference^8^ by sodium dodecyl sulphate polyacrylamide gel electrophoresis (SDS-PAGE) densitometry.

Unmasked staff administered study treatments by intramuscular injection of 0·5 mL on study days 1 and 29. Blood samples were drawn and clinical assessments were done for safety and immunogenicity endpoints before vaccination on days 1 (dose one), 8, 29 (dose two), 36, and 43; a clinical assessment for safety only on day 57 was the last timepoint considered for this interim analysis of the phase 1 cohort, although there will be additional immunogenicity and safety assessments on study days 197 and 365. Subjects were observed in the clinic for 30 minutes after each vaccination and were asked to record any adverse events using paper diary cards during the 7-days after each vaccination. Subjects randomly allocated to a cell-mediated immunity subset (N=12 per 10 µg, 3 µg+CpG1018, and placebo groups) had additional blood collected on days 1 and 43 for isolation of peripheral blood mononuclear cells (PBMCs); these were stored in liquid nitrogen until analysed.

Solicited injection site reactions (pain, tenderness, swelling, induration, erythema) and systemic symptoms (headache, fatigue, malaise, myalgia, arthralgia, nausea, vomiting, and fever defined as oral temperature ≥38°C) were recorded by subjects in a diary card that included intensity, then reported by the investigators; these events were not assessed for causality. Subjects also recorded spontaneously reported adverse events (AEs) for 28 days; the investigator reported these after grading them for intensity and categorizing them as serious or not. The investigator also identified the following AEs of special interest: potential immune-mediated medical conditions (PIMMCs), and AEs of special interest associated with COVID-19. Intensity of AEs was graded 1-4 as follows: 1 or mild (minimal interference with daily activities), 2 or moderate (interferes with but does not prevent daily activities), 3 or severe (prevents daily activities, intervention required), and 4 or very severe (medical intervention required to prevent disability or death). Investigators assessed unsolicited adverse events for causality (related to vaccination or not). AEs were graded according to U.S. Department of Health and Human Services severity grading tables (Food and Drug Administration, Center for Biologics Evaluation and Research [September 2007] and National Institutes of Health, Division of AIDS [version 2.1, July 2017]). A protocol safety review committee regularly reviewed blinded safety data; a Data Safety Monitoring Board monitored unblinded safety data and recommended two formulations for advancing to phase 2.

We measured total anti-SARS-CoV-2 spike (S) IgG using a validated indirect ELISA at Nexelis (Laval, Canada). Purified recombinant SARS-CoV-2 pre-fusion spike (Nexelis) at 1µg/ml in phosphate buffered saline (PBS, Wisent Bioproducts) was adsorbed to 96 well Nunc Maxisorb microplates (Thermo Fischer Scientific) and blocked with 5% skim milk in PBS, containing 0·05% Tween 20. Serial dilutions of test samples and the assay standard plus controls were added in the plates and incubated for 60 minutes at room temperature (15-30°C). After washing, horseradish peroxidase (HRP) enzyme-conjugated goat anti-human IgG-Fc (Jackson ImmunoResearch Laboratories) was added for 60 minutes at room temperature (15-30°C), then washed. Bound secondary antibody was reacted with 3,3′,5,5′-tetramethylbenzidine (TMB) ELISA peroxidase substrate (Bio-Rad Laboratories) and incubated for 30 minutes at room temperature (15-30°C) before the reaction was stopped with 2N H_2_SO_4_. Plates were read at 450 nm with a correction at 620 nm to assess the level of anti-S IgG bound in the microtiter plate. A reference standard on each plate determined the quantity of anti-S IgG in arbitrary units (ELU/mL). Concentrations were transformed to binding antibody units per mL (BAU/mL), based on the WHO International Standard for anti-SARS-CoV-2 immunoglobulin,^9^ using a conversion factor determined during assay validation (1/7·9815). The assay’s cut-off and lower limit of quantitation (LLOQ) was 6·3 BAU/mL.

We measured serum neutralising activity against the Wuhan strain of SARS-CoV-2 in a validated pseudotyped virus neutralisation assay (PNA) that assessed particle entry-inhibition.^10^ Briefly, pseudotyped virus particles containing a luciferase reporter for detection were made from a modified vesicular stomatitis virus (VSVΔG) backbone expressing the full-length spike glycoprotein of SARS-CoV-2 (MN908947, Wuhan-Hu-1) from which the last 19 amino acids of the cytoplasmic tail were removed.^11^ Seven two-fold serial dilutions of heat-inactivated serum samples were prepared in 96-well round-bottom transfer plates (Corning). Pseudotyped virus was added to the serum dilutions at a target working dilution and incubated at 37°C with 5% CO_2_ for 60 ± 5 minutes. Serum-virus complexes were then transferred onto 96 well white flat-bottom plates (Corning), previously seeded overnight with Vero E6 cells (Nexelis) and incubated at 37°C and 5% CO_2_ for 20 ± 2 hours. Following this incubation, luciferase substrate from ONE Glo™ Ex luciferase assay system (Promega) was added to the cells. Plates were then read on a SpectraMax® i3x plate reader (Molecular Devices) to quantify relative luminescence units (RLU), inversely proportional to the level of neutralising antibodies present in the serum. The neutralising titre of a serum sample was calculated as the reciprocal serum dilution corresponding to the 50% neutralisation antibody titre (NT_50_) for that sample; the NT_50_ titres were transformed to international units per mL (IU/mL), based on the WHO international standard for anti-SARS-CoV-2 immunoglobulin, using a conversion factor determined during assay validation (1/1,872). The assay’s cut-off and LLOQ were 5·3 IU/mL (10 as NT_50_) and 5.9 IU/mL, respectively. To benchmark vaccine immunogenicity assessed in BAU/mL and IU/mL, we used a panel of human convalescent serum samples (HCS) collected 14 days after symptom onset from consecutive cases of mild to moderate COVID-19 illness among health care personnel seen as outpatients in Quebec, Canada during mid-2020. We also calculated 80% neutralisation titres (NT_80_); nevertheless, as the PNA was not validated for this measurement, these results are not presented. We used the same PNA assay to measure NT_50_ (reported as titres) against pseudotyped virus particles generated for SARS-CoV-2 variants of concern B.1.315^12^ and P.1^13^. In the absence of positive controls for the variant strains of SARS-CoV-2, we used control sera for the Wuhan-Hu-1 strain.

To assess cellular immunity, we quantified interferon-γ (IFN-γ) and IL-5 producing cells in PBMCs stimulated with SARS-CoV-2 spike peptide pools (vial 1 158 overlapping peptides, vial 2 157 overlapping peptides; JPT Peptide) using a human IFN-γ/IL-5 double-colour ELISpot kit (Cellular Technologies) in a qualified assay. Briefly, activated 96-well plates were coated with anti-human IFN-γ/IL-5 capture antibodies at 2-8°C. Following overnight (>16h) coating, plates were washed with PBS, and stimulation media containing SARS-CoV-2 peptide pool 1 or peptide pool 2 or control media was added to wells, followed by the addition of PBMCs at 2×10^5^ cells/well. After an approximately 44-hour incubation at 37°C±1°C with 5% CO_2_, plates were washed to remove cells from the wells. Anti-human IFN-γ/IL-5 detection solution (containing anti-human IFN-γ fluorescein isothiocyanate [FITC] and anti-human IL-5 [biotin] detection antibodies) was then added to the wells and incubated at room temperature (15-30°C) for 2 h ± 10min to detect IFN-γ and IL-5 cytokine captured on the bottom of the well. Plates were washed, followed by the addition of a tertiary solution (containing FITC-HRP and streptavidin-alkaline phosphatase). Following incubation with the tertiary solution, plates were washed, and blue and red developer solutions were added in sequence (with washes in between), resulting in the appearance of blue (for IL-5) and red (for IFN-γ) spot forming units (SFUs) in proportion to T cell activity. SFUs were counted by an ImmunoSpot CTL Analyzer (using CTL ImmunoCapture Software (v7·0·14·0) and CTL ImmunoSpot Professional DC Analyzer (v7·0·28·2)). Readouts (one per peptide pool for IFN-γ, one per peptide pool for IL-5) were expressed as number of SFU/10^6^ cells and combined as a ratio. The assay’s LLOQ for IFN-γ was 109 SFU/10^6^ cells and for IL-5 was 43 SFU/10^6^ cells.

### Outcome

The primary outcomes were frequency and intensity of solicited injection site and systemic AEs during 7 days after vaccination; frequency, intensity, and relatedness of clinically significant haematological and biochemical measurements at 7 days after each vaccination; frequency, intensity, and relatedness of unsolicited AEs during 28 days after each vaccination; and occurrence of medically-attended AEs, serious AEs, and AEs of special interest during the interim analysis period of 57 days after-first vaccination. The secondary immunogenicity outcomes were anti-S IgG and NT_50_ against Wuhan-1 strain SARS-CoV-2 pseudotyped virus assessed on days 29 and 43 and expressed as geometric mean titre (GMT) or concentration (GMCs, BAU/mL for ELISA, or IU/mL for PNA), geometric mean fold rise (GMFR) from baseline, and percentage of subjects with ≥4-fold increase and ≥10-fold increase from baseline. The exploratory immunogenicity outcomes were cell-mediated immunity to SARS-CoV-2 S protein, measured as the ratio of IFN-γ/IL-5 expressing cells on days 1 and 43 in a random subset of subjects receiving two vaccine formulations (10 µg or 3 µg+CpG1018) or placebo. We also assessed NT_50_ GMTs and the percentage of subjects with a NT_50_ titre ≥4-fold higher than the LLOQ (1:10 titre), against vaccine-heterologous SARS-CoV-2 pseudotyped virus (SARS-CoV-2 variants of concern B.1.351 and P.1) on day 43.

### Statistical Analyses

This Phase 1/2 study (ClinicalTrial.gov NCT04764422) has a two-part selection design with group elimination after the interim analysis. In the first part, 35 subjects per group were randomized across 5 candidate vaccine formulations and a placebo group for a total of 210 subjects. After the interim analysis, two candidates were selected to advance, at which time 250 additional subjects are to be randomized 2:2:1 to the two selected candidate groups and the placebo, respectively. The study was designed to have greater than 90% power to identify the candidate with the highest response as measured by the NT_50_ by ranked GMCs, assuming the true GMC is at least 1·5-fold larger than the second highest candidate group and to provide a preliminary safety evaluation of the candidates. An independent data monitoring committee provided safety oversight.

All statistical tests were two-sided with a significance level of 0·05. All statistical analyses were performed by an independent statistician using SAS version 9·4. All safety assessments took place in the treatment-exposed population, according to the treatment received. All subject-level percentages were supplemented with two-sided 95% confidence intervals computed via the Clopper-Pearson method. The analysis of immunogenicity was performed in the per protocol population, which excludes subjects with protocol deviations that would affect the assessment. Immunogenicity data were descriptively analysed. Geometric mean antibody responses were reported by treatment and time point, accompanied by 95% CIs. The analysis of geometric means excluded subjects who were seropositive at baseline (defined by anti-S IgG >LLOQ as measured by ELISA). Geometric mean fold rises (GMFR) were calculated relative to baseline using the log difference of the paired samples, with corresponding CIs computed via the *t*-distribution, utilizing the antilog transformation to present the ratio. The proportions of subjects with GMFRs of NT_50_ ≥4 and ≥10 from baseline were summarized with 95% CIs. The analysis of immunogenicity relative to baseline included baseline seropositive subjects.

## Results

Between March 22 and April 23, 2021, 210 healthy adults were enrolled and assigned to one of six treatment groups as shown in Figure 1. All received a first dose of vaccine or placebo; two subjects were excluded from receipt of a second dose (one became pregnant, one developed mild urticaria within 30 minutes after dose one); three other subjects missed the day 29 visit and got no second dose. The baseline characteristics are shown by treatment group in table 1; the exposed population was 61% female, had a median age of 36 years (IQR 28, 43) and a median body mass index of 24·07 (IQR 21·30-26·72).

**Figure 1.**
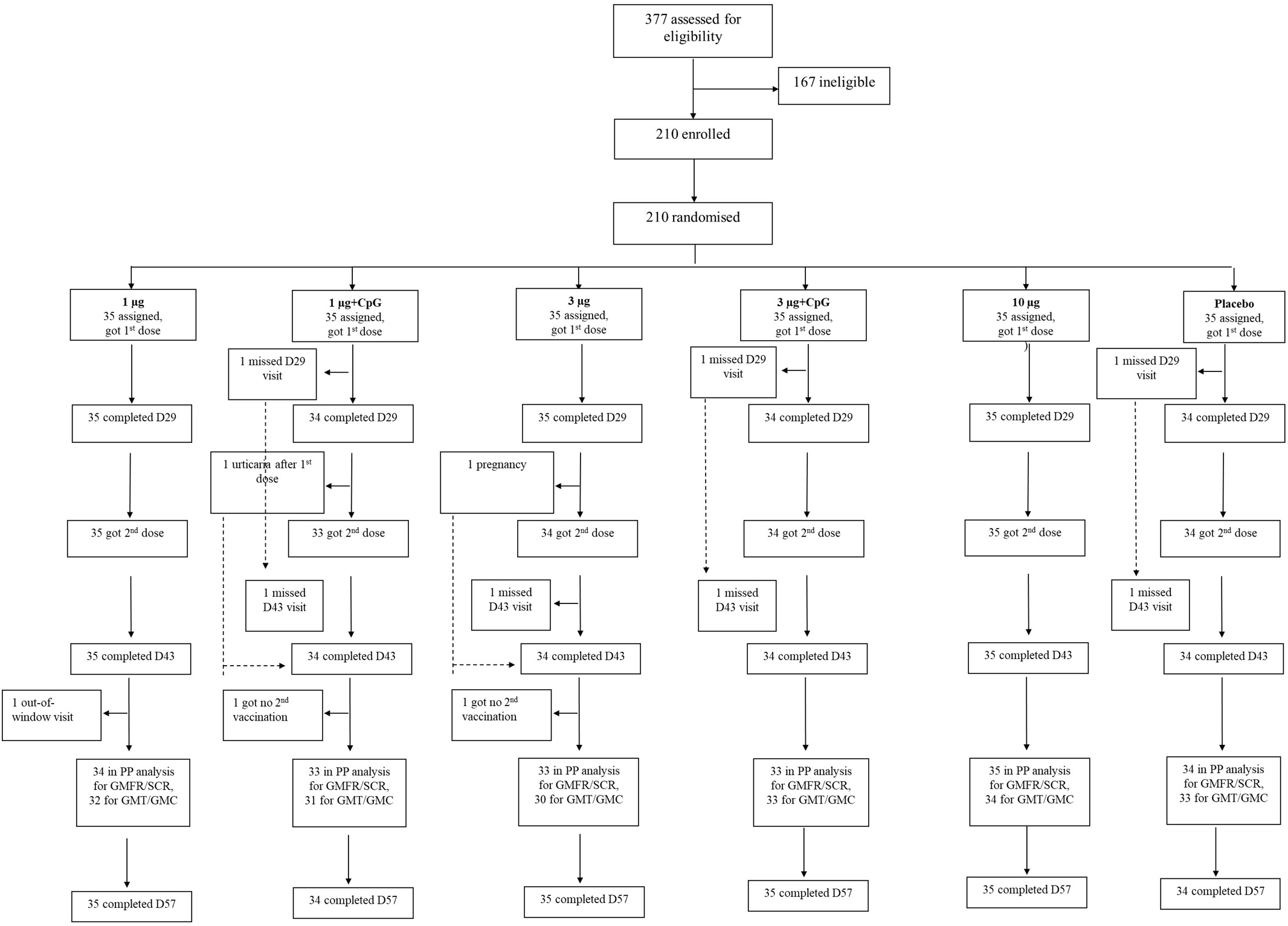
Trial profile

**Table 1.** Baseline characteristics of the exposed population

|  | **1 µg S**<br>**(N = 35)** | **1 µg S+CpG**<br>**(N = 35)** | **3 µg S**<br>**(N = 35)** | **3 µg S+CpG**<br>**(N = 35)** | **10 µg S**<br>**(N = 35)** | **Placebo**<br>**(N = 35)** |
| --- | --- | --- | --- | --- | --- | --- |
| Age, years | 33·0 (26·0-39·0) | 39·0 (32·0-45·0) | 37·0 (29·0-49·0) | 34·0 (25·0-44·0) | 37·0 (31·0-42·0) | 32·0 (27·0-42·0) |
| Sex |  |  |  |  |  |  |
| Male | 14 (40·0%) | 14 (40·0%) | 7 (20·0%) | 15 (42·9%) | 18 (51·4%) | 14 (40·0%) |
| Female | 21 (60·0%) | 21 (60·0%) | 28 (80·0%) | 20 (57·1%) | 17 (48·6%) | 21 (60·0%) |
| Ethnicity |  |  |  |  |  |  |
| Asian | 35 (100%) | 35 (100%) | 35 (100%) | 35 (100%) | 35 (100%) | 35 (100%) |
| Body mass<br>index | 24·59 (20·76-27·85) | 24·85 (21·42-26·33) | 23·95 (21·23-27·96) | 23·95 (21·70-25·92) | 24·52 (21·26-27·68) | 23·12 (21·72-27·22) |

Data are median (q1-q3) or n (%)

All five formulations of NDV-HXP-S were well tolerated with no dose limiting reactogenicity (Table 2). Most solicited injection site and systemic reactogenicity during 7 days after each vaccination was mild and transient with no apparent difference between dose 1 and 2. The most common injection site symptoms (table 2) were pain and tenderness; these were most frequent at the highest dose. The most common systemic symptoms (table 2) were fatigue, headache, and myalgia, generally in less than one-third of subjects. Fever was uncommon. Adverse events occurring during 28 days after vaccination (table 3) and judged by the investigator to be treatment-related were infrequent (<15%) and there was no treatment-related serious adverse event, nor any AE of special interest reported during the 57 day assessment period. Haematology and serum chemistry laboratory readouts were assessed on day 8 following each vaccination; there was no clinically notable finding relative to baseline assessment. The independent data monitoring committee expressed no safety concern.

**Table 2.**
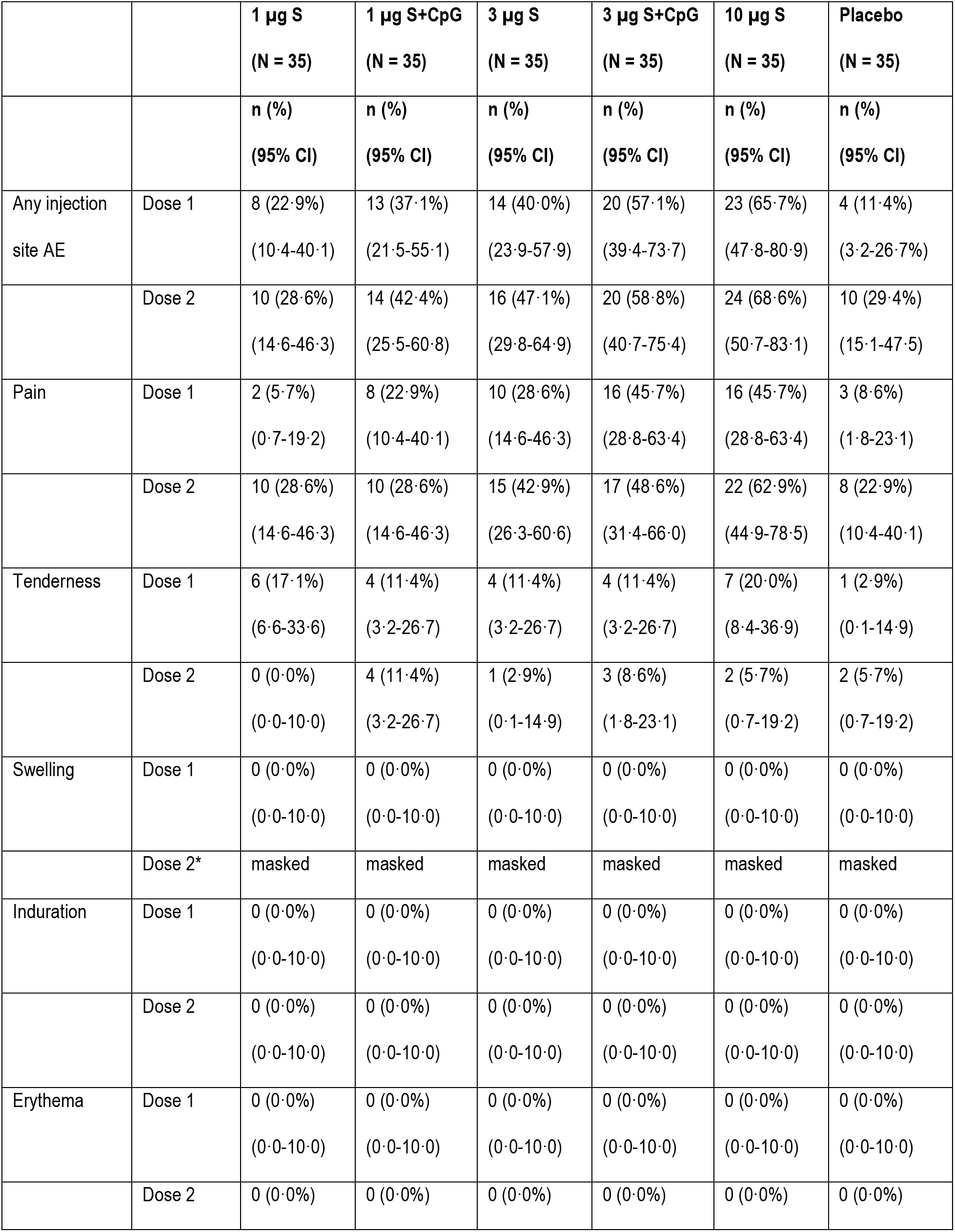
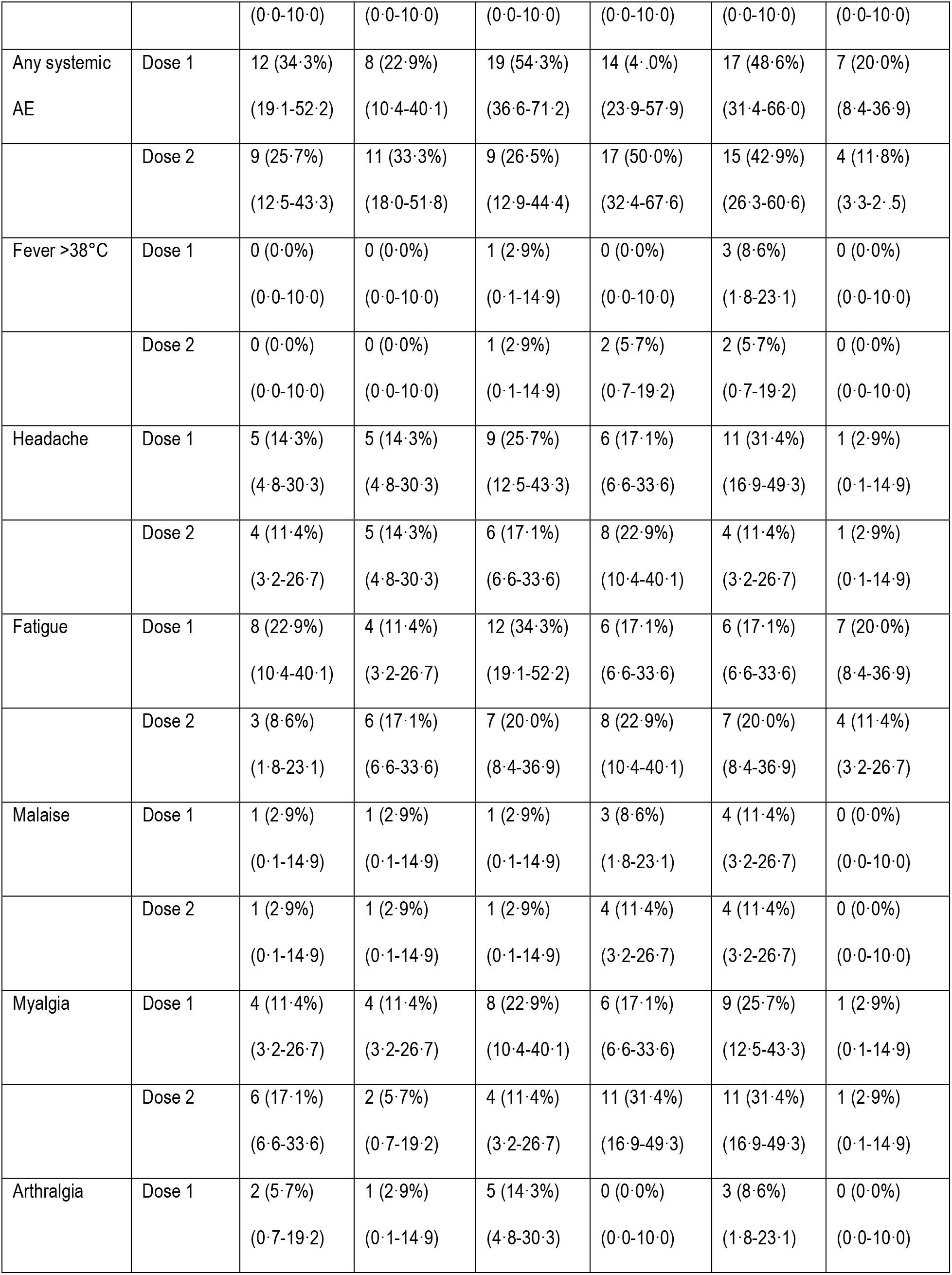
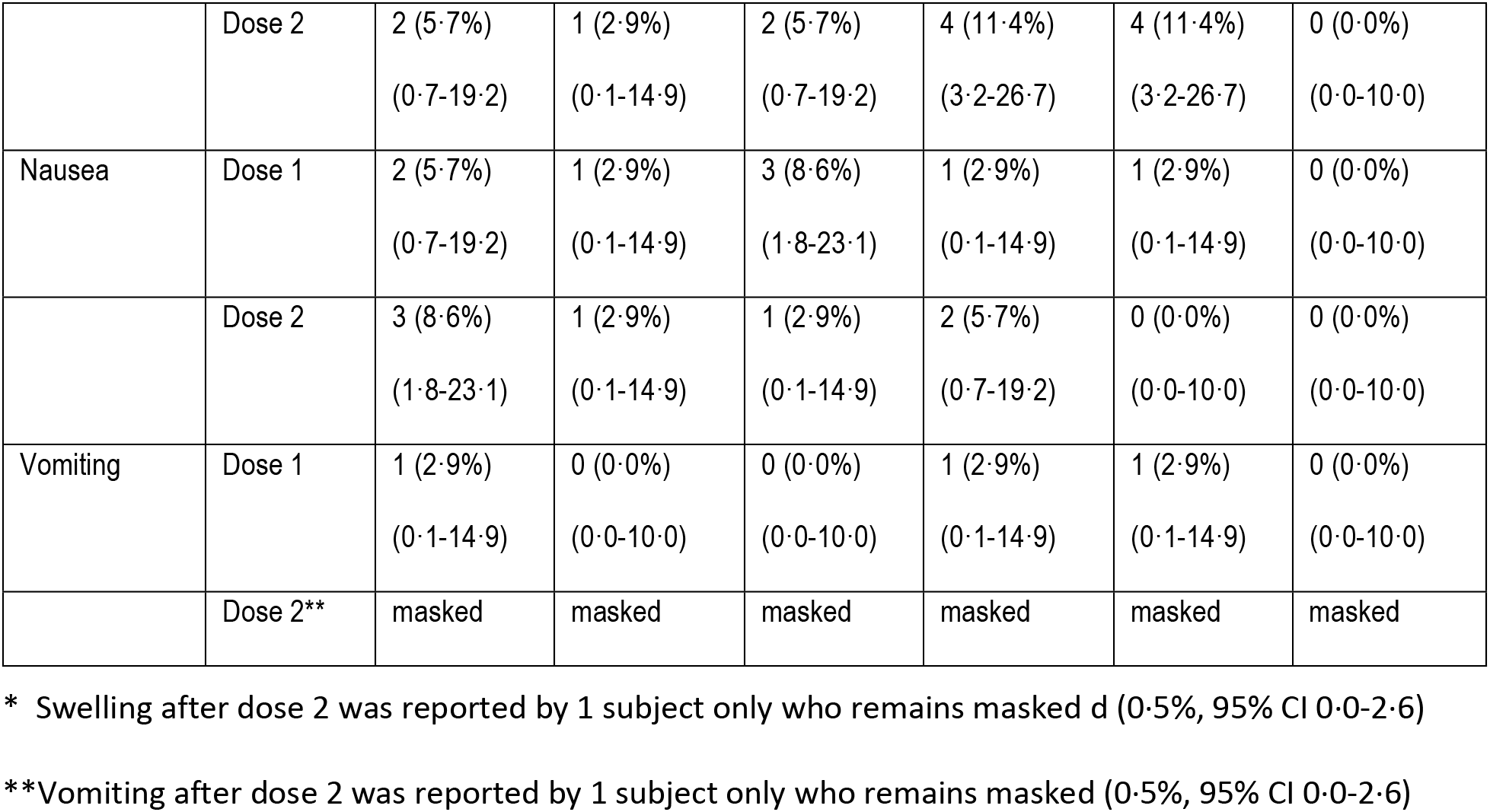
Solicited AEs during 7 days after vaccination

**Table 3.** AEs with onset during 28 days after vaccination

|  |  | **1 µg S**<br>(N = 35) | **1 µg S+CpG**<br>(N = 35) | **3 µg S**<br>(N = 35) | **3 µg S+CpG**<br>(N = 35) | **10 µg S**<br>(N = 35) | **Placebo**<br>(N = 35) |
| --- | --- | --- | --- | --- | --- | --- | --- |
|  |  | n (%)<br>(95% CI) | n (%)<br>(95% CI) | n (%)<br>(95% CI) | n (%)<br>(95% CI) | n (%)<br>(95% CI) | n (%)<br>(95% CI) |
| Any | Dose 1 | 8 (22·9%)<br>(10·4-40·1) | 7 (20·0%)<br>(8·4-36·9) | 15 (42·9%)<br>(26·3-60·6) | 13 (37·1%)<br>(21·5-55·1) | 9 (25·7%)<br>(12·5-43·3) | 6 (17·1%)<br>(6·6-33·6) |
|  | Dose 2 | 7 (20·0%)<br>(8·4-36·9) | 3 (8·6%)<br>(1·8-23·1) | 11 (31·4%)<br>(16·9-49·3) | 10 (28·6%)<br>(14·6-46·3) | 6 (17·1%)<br>(6·6-33·6) | 9 (25·7%)<br>(12·5-43·3) |
| Vaccine-related | Dose 1 | 2 (5·7%)<br>(0·7-19·2) | 3 (8·6%)<br>(1·8-23·1) | 3 (8·6%)<br>(1·8-23·1) | 5 (14·3%)<br>(4·8-30·3) | 3 (8·6%)<br>(1·8-23·1) | 0 (0·0%)<br>(0·0-10·0) |
|  | Dose 2 | 0 (0·0%)<br>(0·0-10·0) | 1 (2·9%)<br>(0·1-14·9) | 2 (5·7%)<br>(0·7-19·2) | 2 (5·7%)<br>(0·7-19·2) | 2 (5·7%)<br>(0·7-19·2) | 1 (2·9%)<br>(0·1-14·9) |
| Serious | Dose 1* | masked | masked | masked | masked | masked | masked |
|  | Dose 2 | 0 (0·0%)<br>(0·0-10·0) | 0 (0·0%)<br>(0·0-10·0) | 1 (2·9%)<br>(0·1-14·9) | 0 (0·0%)<br>(0·0-10·0) | 1 (2·9%)<br>(0·1-14·9) | 1 (2·9%)<br>(0·1-14·9) |
| Serious vaccine-related | Dose 1 | 0 (0·0%)<br>(0·0-10·0) | 0 (0·0%)<br>(0·0-10·0) | 0 (0·0%)<br>(0·0-10·0) | 0 (0·0%)<br>(0·0-10·0) | 0 (0·0%)<br>(0·0-10·0) | 0 (0·0%)<br>(0·0-10·0) |
|  | Dose 1 | 0 (0·0%)<br>(0·0-10·0) | 0 (0·0%)<br>(0·0-10·0) | 0 (0·0%)<br>(0·0-10·0) | 0 (0·0%)<br>(0·0-10·0) | 0 (0·0%)<br>(0·0-10·0) | 0 (0·0%)<br>(0·0-10·0) |

\* Serious AE was reported by one subject only who remains masked (0·5%, 95% CI 0·0-2·6)

Two doses of NDV-HXP-S were immunogenic in a formulation and dose dependent manner within the per protocol population. Induction of anti-S IgG was modest following dose one but there was a marked anamnestic response observed 14 days after vaccine dose two (figure 2A). Seronegative individuals in the vaccine groups responded 28 days after first vaccination with GMCs of anti-S IgG between 7·79 (1 µg) and 20·93 (10 µg) BAU/mL, with a ≥4-fold increase in 34·3-71·4%. The second dose considerably increased anti-S-IgG antibody responses after 14 days to GMCs between 151·78 (1 µg) and 479·83 (10 µg) BAU/mL. All individuals in every vaccine group had a ≥ 4-fold increase over baseline after the second dose; all individuals in the 10 µg and 3 ug+CpG1018 groups had a ≥10-fold increase, as did >90% of vaccinees in the other three vaccine groups (figure 2C). Notably, the adjuvant effect of CpG was limited after two vaccine doses (table 4): the 1 µg group had a GMC of 151·78 BAU/mL (95% CI 108·99-211·37) while the 1 µg+CpG1018 group had a GMC of 199·08 BAU/mL (95% CI 140·25-282·57). Among recipients of the 3 µg dose, the GMC group difference appeared to be greater: 228·07 BAU/mL (no adjuvant, 95% CI 154·22-337·27) in contrast to 356·83 BAU/mL (CpG1018, 95% CI (265·89-478·88). GMCs of anti-S IgG among the vaccine groups on day 43 exceeded the GMC of the HCS panel (N=29, 72·93 95% CI 33.00-161.14) by 2-6-fold (table 4).

**Figure 2.**
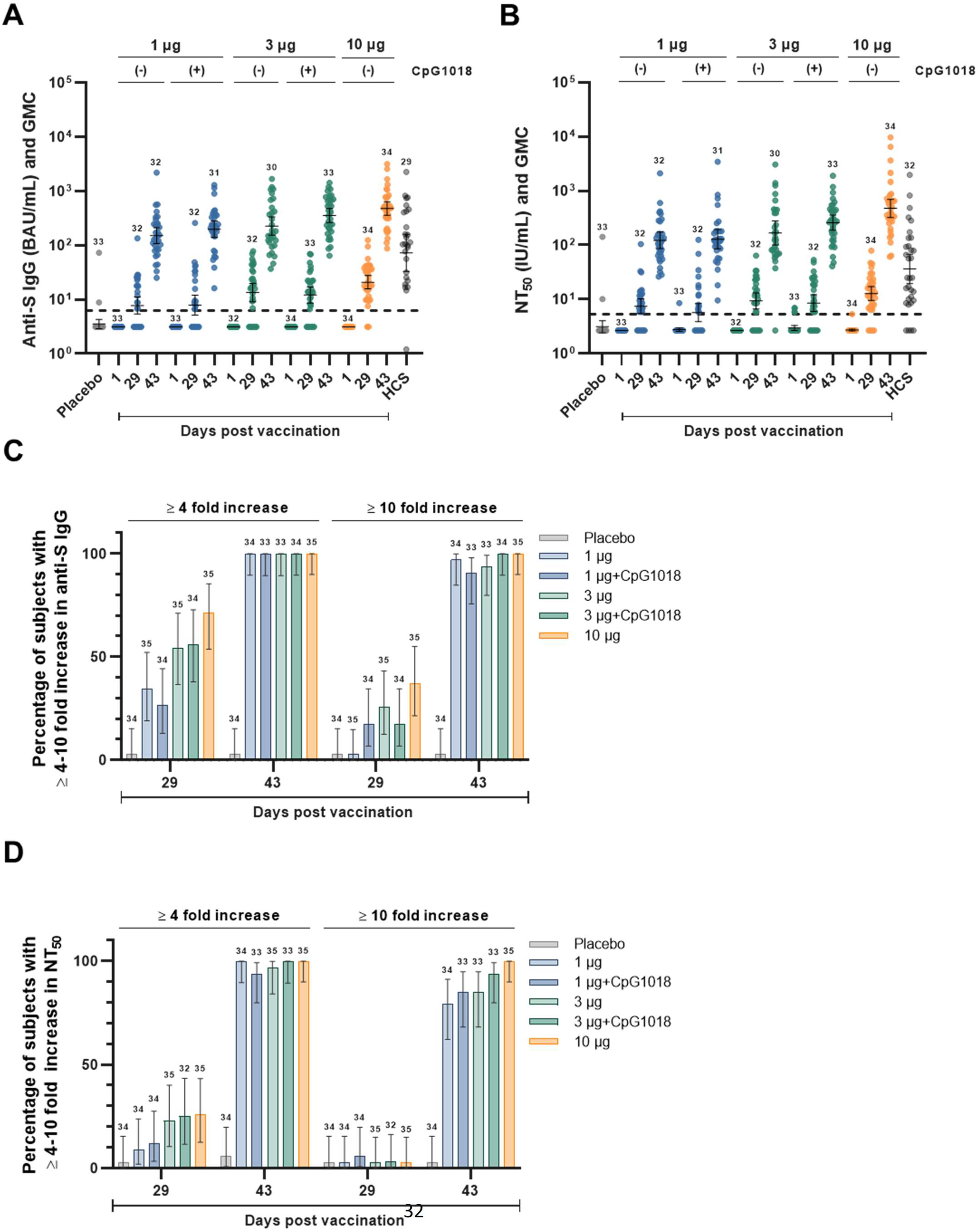
Distribution and GMC of anti-S IgG (BAU/mL) in placebo, vaccine groups and HCS controls (A), distribution and GMC of NT_50_ by PNA (IU/mL) in placebo, vaccine groups, and HCS controls (B), percentage of subjects with ≥4-10-fold increase in anti-S IgG (C), and percentage of subjects with ≥4-10-fold increase in NT_50_ by PNA (D); numbers above data denote number of per-protocol subjects contributing data

**Table 4.** GMCs of anti-S IgG (BAU/mL) and NT_50_ by PNA (IU/mL) on day 43 and GMC ratios, vaccine to HCS panel

|  |  | **1 µg S** | **1 µg S+CpG** | **3 µg S** | **3 µg S+CpG** | **10 µg S** |
| --- | --- | --- | --- | --- | --- | --- |
| Anti-S IgG BAU/mL, | GMC | 151·78 | 199·08 | 228·07 | 356·83 | 479·83 |
|  | 95% CI | (108·99-211·37) | (140·25-282·57) | (154·22-337·27) | (265·89-478·88) | (360·19-639·20) |
|  | GMC ratio,<br>vaccine to<br>HCS panel | 2·08 | 2·73 | 3·13 | 4·89 | 6·58 |
|  | 95% CI | (0·89-4·87) | (1·16-6·43) | (1·31-7·48) | (2·12-11·31) | (2·85-15·18) |
| NT50 by PNA | GMC | 122·23 | 127·92 | 166·54 | 257·70 | 474·35 |
|  | 95% CI | (86·40-172·91) | (85·08-192·34) | (100·19-276·81) | (187·01-355·11) | (320·90-701·19) |
|  | GMC ratio,<br>vaccine to<br>HCS panel | 3·37 | 3·52 | 4·59 | 7·10 | 13·07 |
|  | 95% CI | (1·67-6·81) | (1·69-7·34) | (2·08-10·10) | (3·55-14·20) | (6·33-26·99) |

Functional antibody responses were assessed by PNA. Low NT_50_ GMCs were detected in all vaccine groups after the first vaccination (between 7·49 IU/mL and 12·82 IU/mL) with ≥4-fold rises in 8·8% to 25·7% of the vaccine groups (figure 2B, 2D). The second vaccine dose strongly boosted neutralisation GMCs to between 122·23 IU/mL (1 µg, 95% CI 86·40-172·91) and 474·35 IU/mL (10 µg, 95% CI 320·90-701·19), with a ≥4-fold increase over baseline in 93·9% to 100% of vaccine groups and a ≥10-fold rise in most individuals (100% in the 10 µg group, and between 79·4% and 93·9% in the remaining groups). The differences in post-second dose GMCs between the unadjuvanted and adjuvanted 1 µg and 3 µg groups were uncertain: 1 µg, 122·23 IU/mL (95% CI 86·40-172·91) versus 1 µg+CpG1018, 127·92 IU/mL (95% CI 85·08-192·34); 3 µg, 166·54 IU/m: (95% CI 100·19-276·81) versus 3 µg+CpG1018 257·70 IU/mL (95% CI 187·01-355·11).

Based on the vaccine-homologous binding and neutralising antibody responses, there was a clear ranking of immunogenicity with the 10 µg formulation performing best followed by the 3 µg+CpG1018, 3 µg, 1 µg+CpG1018 and 1 µg formulations. The induction of humoral immunity was strong with post-boost GMFRs relative to baseline of 48-fold (1 µg) to 152-fold (10 µg) for anti-S IgG and 46-fold (1 µg) to 174-fold (10 µg) for NT_50_ antibodies (figure 3). GMCs of NT_50_ by PNA among the vaccine groups on day 43 exceeded the GMC of the HCS panel (N=32, 36·30 95% CI 19·43-67·79) by 3-13-fold depending on the vaccine formulation (table 4).

**Figure 3.**
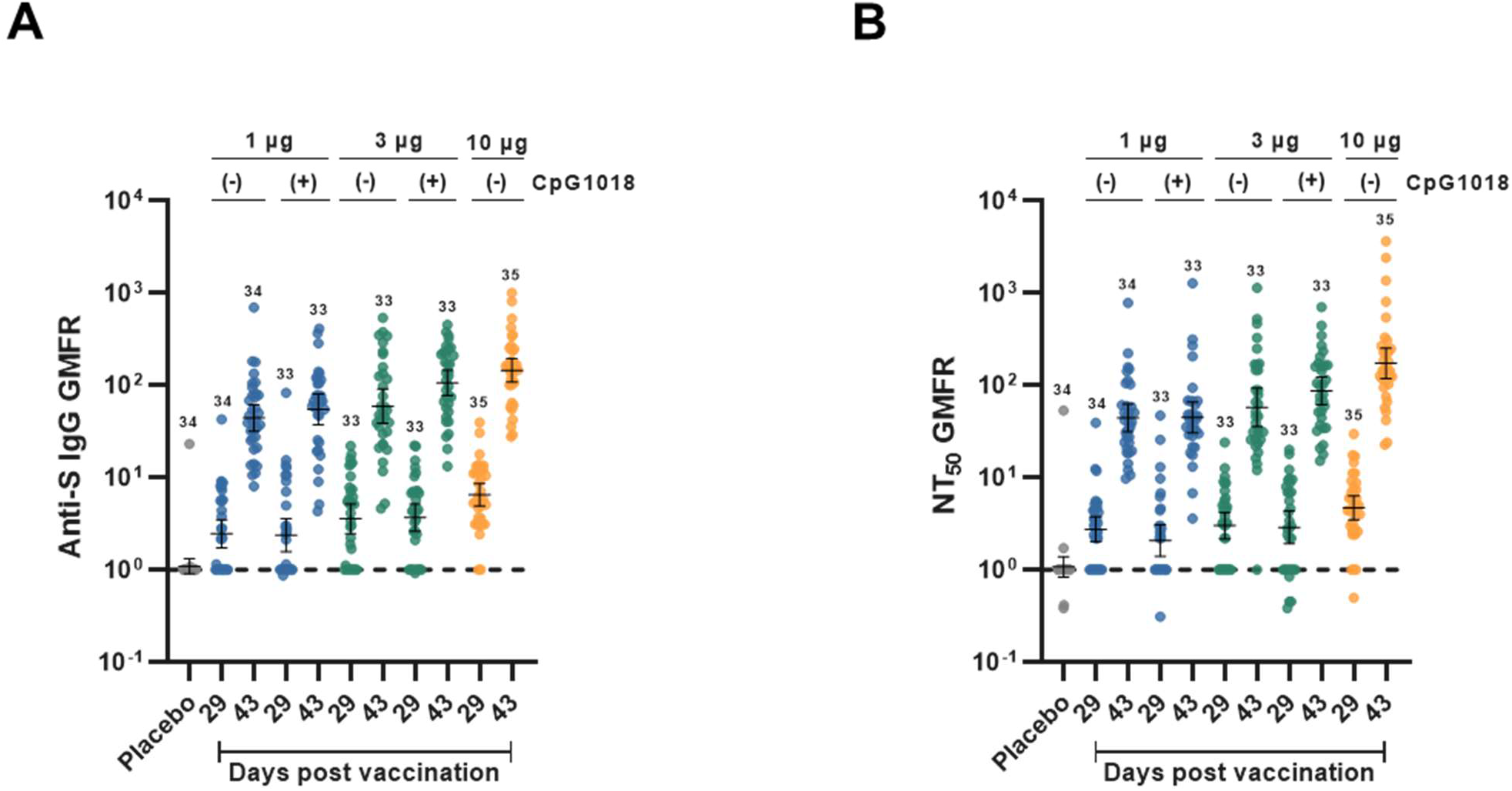
Distribution and GMFR of fold rise in anti-S IgG from baseline (A), distribution and GMFR of fold rise in NT_50_ by PNA from baseline (B); numbers above data denote number of per-protocol subjects contributing data

Additionally, neutralisation of variant viruses was assessed by PNA on day 43. The proportion of subjects attaining a day 43 NT_50_ titre ≥40 increased with higher doses of antigen but the incremental changes in GMT were small. Reduction in neutralising potency, relative to anti-Wuhan neutralising potency, was modest for the P.1 variant (2·8 to 5·3-fold) but greater for the B·1·351 variant (7·41 to 20·43-fold) (figure 4, table 5). In groups receiving 3 µg or 10 µg antigen doses, the proportion attaining a NT_50_ titre ≥40 ranged from 80·0% to 94·9% against P.1 and from 43·3% to 58·5% against B.1.351 (table 5). Finally, we also explored T cell responses to determine if the vaccine induced primarily a type 1 (T_H_1) or type 2 (T_H_2) T-helper cell response. In the small subset of subjects evaluated 14 days after a second dose, the IFN-γ/IL-5 ratio was strongly skewed to a T_H_1 response relative to pre-vaccination baseline (figure 5), suggesting the vaccine induced T cell memory capable of an antiviral response.

**Figure 4.**
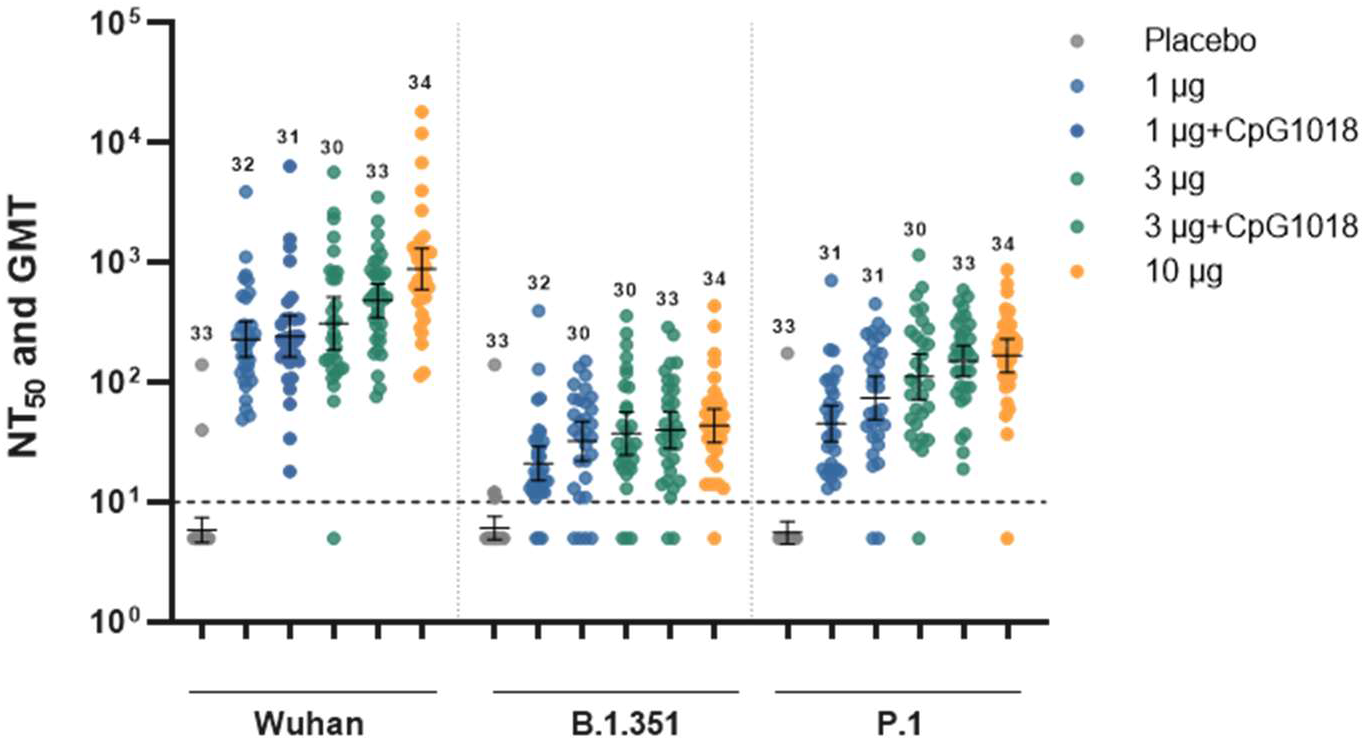
Distribution and GMT of NT_50_ by PNA against variants of concern (day 43); numbers above data denote number of per-protocol subjects contributing data

**Table 5.** GMT and percentage of subjects with a ≥4-fold rise from baseline (day 43) for NT_50_ by PNA against Wuhan strain and variants of concern

|  |  | 1 µg S | 1 µg S+CpG | 3 µg S | 3 µg S+CpG | 10 µg S | Placebo |
| --- | --- | --- | --- | --- | --- | --- | --- |
| Wuhan | GMT | n = 32<br>228·81<br>(161·74-323·69) | n = 31<br>239·47<br>(159·27-360·06) | n = 30<br>311·76<br>(187·56-518·18) | n = 33<br>482·42<br>(350·08-664·77) | n = 34<br>887·99<br>(600·72-1,312·62) | n = 33<br>5·89<br>(4·64-7·48) |
| | $\geq 4$-fold rise | N = 34<br>34 (100%)<br>(89·7-100) | N = 33<br>31 (93·9%)<br>(79·8-99·3) | N = 33<br>32 (97·0%)<br>(84·2-99·9) | N = 33<br>33 (100%)<br>(89·4-100) | N = 35<br>35 (100%)<br>(90·0-100) | N = 34<br>2 (5·9%)<br>(0·7-19·7) |
| P·1 | GMT | n = 31<br>45·33<br>(32·18-63·85) | n = 31<br>74·06<br>(48·99-111·97) | n = 30<br>111·28<br>(72·06-171·83) | n = 33<br>150·59<br>(111·15-204·03) | n = 34<br>167·14<br>(120·63-231·58) | n = 33<br>5·57<br>(4·47-6·94) |
| | $\geq 4$-fold rise | N = 31<br>15 (48·4%)<br>(30·2 –<br>66·9) | N = 31<br>29 (74·2%)<br>(55/4 – 88·1) | N = 30<br>24 (80·0%)<br>(61·4 –<br>92·3) | N = 33<br>29 (87·9%)<br>(71·8 – 96·6) | N = 34<br>(32 (94·9%)<br>(80·3 – 99·3) | N = 33<br>1 (3·0%)<br>(0·1 – 5·8) |
| B·1·351 | GMT | n = 32<br>21·00<br>(15·12-29·18) | n = 30<br>32·34<br>(22·13-47·24) | n = 30<br>37·43<br>(24·77-56·56) | n = 33<br>40·07<br>(28·09-57·17) | n = 34<br>43·47<br>(31·55-59·89) | n = 33<br>6·12<br>(4·90-7·64) |
| | $\geq 4$-fold rise | N = 32<br>5 (15·6%)<br>(5·3 –<br>32·8) | N = 40<br>15 (50·0%)<br>(31·3 -68·7) | N = 30<br>(13<br>(43·3%)<br>(30·8 –<br>66·5) | N = 33<br>16 (48·5%)<br>(30·8 – 66·5) | N = 34<br>20 (58·8%)<br>(40·7 – 75·4) | N = 33<br>1 (3·0%)<br>(0·1 – 15·8) |

**Figure 5.**
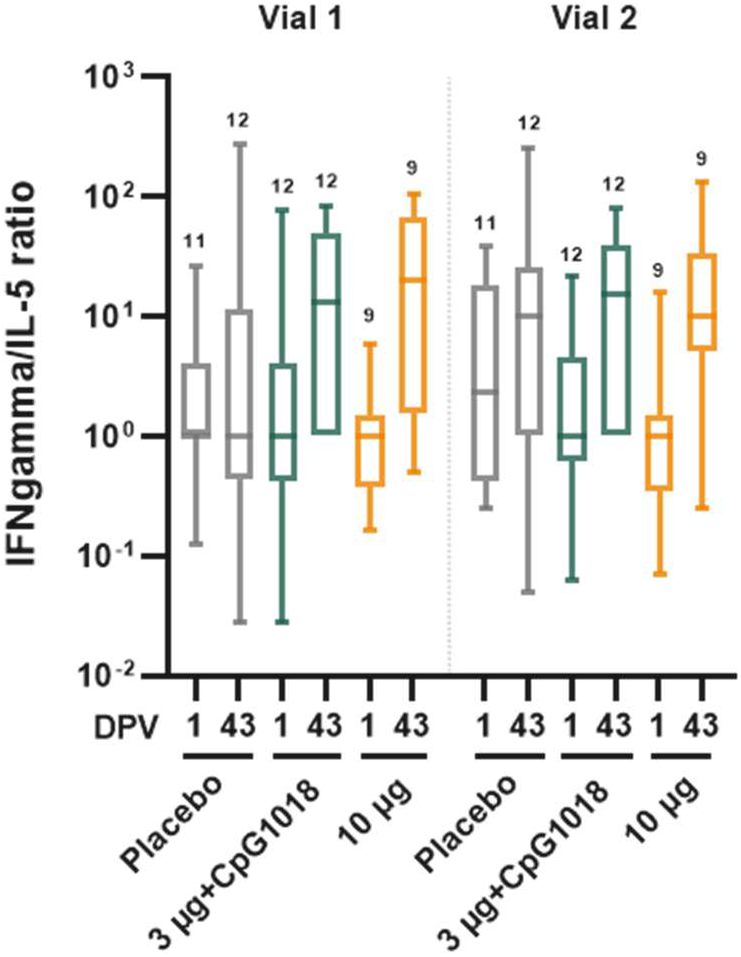
Box plot of IFN-γ /IL-5 ratios (ELISpot 2-colour assay); numbers above data denote number of per-protocol subjects contributing data

## Discussion

Current production capacity cannot satisfy the global demand for COVID-19 vaccines^1^ and vaccine distribution is inequitable with most vaccines acquired and used by high income countries while LMICs have limited access. Local production of COVID-19 vaccines in LMICs would increase global availability and reduce dependence of countries producing these vaccines on international vaccine supply. Here we demonstrated for the first time that an engineered inactivated NDV-based vaccine expressing a second-generation stabilized SARS-CoV-2 spike protein^5^, produced in eggs in an existing influenza virus production facility at GPO in Thailand, shows an acceptable reactogenicity and safety profile in humans and has immunogenicity that supports its potential clinical benefit. We evaluated a range of vaccine doses (1 µg, 3 µg, 10 µg) having potency quantified as µg of virus envelope-anchored SARS-CoV-2 spike protein; the low and medium antigen doses were evaluated in formulations with and without the TLR-9 agonist CpG1018 as a vaccine adjuvant. Over 28 days after each vaccine dose, all formulations were very well-tolerated with little solicited injection site or systemic reactogenicity aside from mild injection site pain and tenderness. There was no safety signal issuing from this early interim analysis of the clinical trial. Moreover, the vaccine was strongly immunogenic in a formulation and dose dependent manner, inducing levels of vaccine-homologous anti-S IgG and virus neutralising antibodies that exceeded by several fold the levels measured in 14-day convalescent sera from consecutive cases of health care workers with mild to moderate COVID-19 illness in 2020. Notably, the adjuvant benefit as measured by enhanced induction of humoral immunity was uncertain, as the small sample size limited precision. On the other hand, the vaccine at all dose levels elicited neutralising antibodies against two variants of concern, B.1.351 and P.1. While neutralising antibody titres decreased modestly against P.1 and more markedly against B.1.351, this was expected and in the range observed with sera from recipients of the mRNA vaccines BNT162b2 and mRNA-1273.^14-17^ The degree of reduction in neutralisation is dependent on the assay used and can be especially dramatic with pseudotyped particle inhibition assays as used in this study.^14,17^ Notably, the B.1.351 variant is currently regarded as a worst case example of immune evasion; accordingly, the NDV-HXP-S would be predicted to have neutralising activity against the now prevalent delta (B.1.617.2) variant. The T cell response assessed showed a bias towards a T_H_1 response in both evaluated dose groups, alleviating concerns about enhanced disease associated with a T_H_2 response (as observed with SARS-CoV-1 in some animal models^18^). These initial data, while sparse, suggest the vaccine-induced T cell memory capable of an antiviral response.

The study has several limitations. The sample size per treatment group was small, limiting precision, and assessments were restricted to 43 days for immunogenicity and 57 days for reactogenicity and safety, narrowing our perspective to acute outcomes only. These are inherent problems of phase 1 trials and interim analyses in a pandemic response setting. Nevertheless, as clinical trials with similar vaccines are underway in Vietnam (NCT04830800) and Brazil (NCT04993209), we determined that publication of early data is a priority. The study had strengths as well. The vaccine construct is a novel platform expressing a second-generation pre-fusion stabilized S protein in a membrane-bound trimeric conformation. We hypothesize that these characteristics contribute to the vaccine’s notable immunogenicity, even without the CpG1018 adjuvant. The anti-S ELISA and PNA used to assess vaccine-homologous NT_50_ potency were validated and results are expressed in International Units^9^ for future comparisons. The induction of anti-S binding and neutralising antibodies was contrasted with mean levels in human convalescent serum and found to be superior, especially in the mid- and high-dose groups. However, the neutralisation assay is not a live virus assay; therefore, it is presently uncertain how our functional antibody readouts can be used to benchmark against live virus neutralising antibody levels induced by authorized or licensed vaccines. Correlation between neutralising antibody titres and vaccine efficacy and individual protection has recently been shown; work is ongoing to integrate these data into this analytic framework.^19-21^ On the other hand, mean vaccine anti-S IgG ELISA responses normalized by the mean in convalescent sera as proposed by Earle and others^19^ suggest that the NDV-HXP-S vaccine will afford important clinical benefit. Furthermore, we plan to conduct a clinical trial in which the NDV-HXP-S vaccine will be contrasted to an authorized comparator vaccine to generate relative immunogenicity evidence that may be predictive of clinical benefit.

In summary, we show that the inactivated NDV-HXP-S vaccine candidate has an acceptable safety profile and is highly immunogenic. This vaccine can be produced at low cost in any facility designed for production of inactivated influenza virus vaccine; such facilities are present in a number of LMICs.^2^ Based on these results, and acknowledging the imperative to maximize output of vaccine doses from the manufacturing facility, the 3 µg and 3 µg+CpG1018 formulations were selected for further assessment in the phase 2 stage of the ongoing clinical trial.

## Data Availability

Data will be available as appropiate.

## Contributors

PPit and SLaw verified the underlying data reported herein. All authors had full access to all the data in the study. Individual author roles are reported using CRediT: Conceptualisation, PPit, SLam, LDM, RSch, AGS, PPal, FK, KP, PW, and BLI; Data curation, PPit and SLaw; Formal analysis, Slaw, LDM, JMC, and JT; Funding acquisition, PN, SSur, RR, NS, AGS, PPal, FK, KP, PW, and BLI; Investigation, PPit, VL, SM, SKam, CS, WS, YL, WP, FA, LG, SKha, SM, IM, BP, SSla, and STra; Methodology, PPit, VL, SM, PN, SSur, SPra, SPuk, SLam, LDM, RR, WS, YL, FA, LG, RK, SKha, ML, IM, BP, RSin, NS, SThe, STra, TV, JAW, FK, and RH, Project administration, PPit, PN, SSur, RSch, YS, NS, NT, TV, and PW; Resources, PPit, SPra, SPuk, WS, CLH, ML, JSM, RSin, SThe, JAW, AGS, PPal, FK, and RH; Supervision, PPit, SPra, RR, RSch, LG, RK, ML, JAW, AGS, PPal, FK, KP, PW, and BLI; Validation, SLaw, SPra, SPuk, LG, RK, SKha, RSin, SThe, and STra; Visualisation, LDM, JMC, JT, and BLI; Writing–original draft, LDM, RR, FK, and BLI; Writing–review & editing, PPit, VL, SLaw, SM, SKam, CS, SSur, SPuk, SLam, WS, JMC, RSch, FA, LG, CLH, RK, SKha, ML, JSM, IM, JT, STra, JAW, PW, and RH.

## Data sharing

Under review, to be decided by the sponsor prior to publication.

## Acknowledgements

Activities at PATH and Mount Sinai were supported, in part, by the Bill & Melinda Gates Foundation [INV-021239]. Under the grant conditions of the Foundation, a Creative Commons Attribution 4.0 Generic License has already been assigned to the Author Accepted Manuscript version that might arise from this submission. The findings and conclusions contained within this manuscript are those of the authors and do not necessarily reflect positions or policies of the Bill & Melinda Gates Foundation. The salary of PP was partially funded by NIH (Centers of Excellence for influenza Research and Response, CEIRR,75N93021C00014), NIAID grant (P01 AI097092-07), NIAID grant (R01 AI145870-03), by the NIH Collaborative Influenza Vaccine Innovation Centers (CIVICs) contract 75N93019C00051 and a grant from an anonymous philanthropist to Mount Sinai. Design and generation of reagents used in this project in the Krammer laboratory were supported by CEIRR (75N93021C00014) and CIVIC (75N93019C00051). Research and development activities in Thailand were funded by the Government Pharmaceutical Organization Thailand), the National Vaccine Institute (Thailand), and the National Research Council (Thailand). Testing of clinical trial specimens at Nexelis was supported by the Global Health Vaccine Accelerator Platform of the Bill & Melinda Gates Foundation. The authors thank Dr. Randy Albrecht for management of import/export of Newcastle disease virus vectors at the Icahn School of Medicine at Mount Sinai. We are grateful to Dr. Nina Bardwaj for programmatic and scientific oversight of the Virus and Cell Therapy Laboratory at the Icahn School of Medicine at Mount Sinai, which produced the NDV-HXP-S master virus seed.

